# Light Chain Neurofilament to HDGFL2 cryptic peptide ratio as a fluid biomarker to monitor TDP-43 dysfunction in ALS and FTD

**DOI:** 10.64898/2025.12.30.25343222

**Authors:** Irene Santos-García, Katherine E. Irwin, Patricia Garay-Albizuri, Fermín Moreno-Izco, Javier Ruiz-Martínez, Adolfo López de Munain, Jonathan P. Ling, Philip C. Wong, Lorea Blázquez

**Author notes:** Correspondence to: Lorea Blázquez, Irene Santos-García.

## Abstract

TDP-43 proteinopathy is a neuropathological hallmark of nearly all amyotrophic lateral sclerosis (ALS) and approximately half of frontotemporal dementia (FTD) cases. Nuclear loss of TDP-43 leads to widespread RNA misprocessing, such as the inclusion of cryptic exons that are no longer repressed by TDP-43. Notably, in-frame cryptic exons encode novel cryptic peptides that can be detected in biofluids, including that found in the *HDGFL2* transcript. Here, we quantified HDGFL2 cryptic peptide and neurofilament light chain (NfL) in paired cerebrospinal fluid (CSF) and plasma samples from ALS and FTD patients. Cryptic HDGFL2 peptide was detected in the CSF of ALS patients, whereas no significant differences were observed between genetic and behavioral FTD subgroups. In contrast, NfL levels were elevated in both ALS and FTD, although this biomarker does not reflect TDP-43 pathology. Notably, NfL:HDGFL2 cryptic peptide ratio outperformed either marker alone in discriminating ALS and FTD cases from controls, achieving high specificity. Moreover, this ratio correlated with disease progression in ALS, suggesting added prognostic value. Collectively, our findings support the NfL:HDGFL2 cryptic peptide ratio as a promising fluid biomarker that integrates neurodegeneration with TDP-43 dysfunction, potentially improving diagnostic accuracy, disease stratification, and longitudinal monitoring in TDP-43–associated neurodegenerative disorders.

## INTRODUCTION

Identifying reliable biomarkers for neurodegenerative disorders has become a crucial area of research, not only for diagnostic and prognostic purposes, but also to facilitate patient recruitment and stratification in clinical trials. Some molecules, such as neurofilament light chain (NfL), are used as general neurodegeneration biomarkers, and appear elevated in patients with several neurodegenerative diseases, including Amyotrophic Lateral Sclerosis (ALS) or Frontotemporal dementia (FTD)^1^. However, no specific fluid biomarkers are available for ALS and FTD^23^. A central pathological hallmark of the ALS-FTD spectrum is the nuclear depletion and cytoplasmic aggregation of the RNA-binding protein TDP-43 ^4^. This dysregulation leads to the loss of TDP-43 repression of cryptic splicing and the inclusion of non-conserved cryptic exons (CE) ^5–7^. Recent investigations have highlighted the potential of cryptic peptides derived from cryptic exons as promising candidates for biomarker development ^8^. One example is HDGFL2 cryptic peptide, which is elevated in CSF from patients with sporadic ALS (sALS) and presymptomatic and symptomatic *C9orf72* carriers ^9^. Other authors have also confirmed elevated levels of HDGFL2 cryptic peptide in *post-mortem* tissue samples positive for TDP-43 proteinopathy ^10^. Results from both studies raise the possibility of using HDGFL2 as a biomarker for diagnosing and monitoring these conditions. However, these results have not been validated in independent cohorts or in patients with other clinical or genetic diagnoses, and data is also missing about their sensitivity and specificity compared to other biomarkers. The present study aims to elucidate the potential of HDGFL2 cryptic peptide and its combination with another neurodegenerative-associated molecule, NfL, as a potential biomarker for TDP-43 proteinopathies. For that, a comprehensive analysis of both biomarkers was performed in paired CSF and plasma samples and associated with clinical outcomes. Our results indicate that a combined biomarker strategy based on HDGFL2 cryptic peptide and NfL enhances diagnostic accuracy between both diseases, which may facilitate early intervention and ultimately improve the quality of life of ALS-FTD patients.

## MATERIALS & METHODS

### Patient classification and biofluid selection

The present single-centre retrospective study involved 85 individuals from the Donostia University Hospital (DUH) (Table S1) with an enrollment period 2012-2024 including: control (n=9), amyotrophic lateral sclerosis (ALS, n=29), frontotemporal dementia (FTD, n=33) and Parkinson’s disease carriers of R1441G mutation in *LRRK2* gene (n=16)^11^. The inclusion criteria for control individuals were the absence of cognitive and motor symptoms, coupled with CSF levels of amyloid-β, Tau and pTau-181 non-suggestive of a neurodegenerative process. Patients belonging to the ALS group include n=23 patients with sporadic ALS (sALS), and several genetic cases including *TARDBP* missense variant c.1055A>G (n=1), *ATP13A2* missense variant c.3380C>T (n=1) and *ARPP21* missense variant c.1586C>T (n=4), recently identified as a risk gene for familial ALS^12^. Individuals belonging to the FTD spectrum comprised the largest and most heterogeneous group. It includes participants with a clinical diagnosis of behavioral FTD (bvFTD) with no genetic cause (n=17), and a second group entitled as genetic FTD (gFTD) which includes *PGRN* c.709-1G>A splice acceptor variant restricted to the Basque Country (n=14)^13,14^, *C9Orf72* (n=1), *TBK1* (n=1) and *VCP* (n=1), all of them suspected of TDP-43 pathology. CSF and plasma samples from the DUH cohort were collected at the same time point and were provided by the Basque Biobank. Samples available for this study were determined in collaboration with the clinical team of the Neurology service of DUH, who were in charge of diagnostic, biofluid collection, and clinical follow-up. Disease progression rate data from ALS patients were included to correlate with biomarker levels. It is calculated between the maximum ALSFRS-R score and the ALSFRS-R score at diagnosis, divided by disease duration (in months)^15,16^. Neither sex nor gender was factored into sample selection to prioritize the number of paired samples acquired, but statistical analyses were performed to evaluate biomarker differences between males and females.

### HDGFL2 cryptic peptide quantification

MSD ELISA technology, developed as previously described ^9^ was applied to measure HDGFL2 cryptic peptide levels in biofluid samples from DUH cohort. In brief, an electrochemiluminescence immunoassay was conducted on MSD MULTI-ARRAY 96-well sector plates using 10µg/µl per well of capture TC1HDG antibody against the cryptic peptide epitope in HDGFL2. CSF and plasma samples were assayed in duplicates of 50µl diluted in 50µl of 1% BSA in PBS (CSF) or MSD Diluent 57 (plasma) to avoid hook effect and fit the assay signal into the linearity range. The standard curve was constructed using a concentration series of purified cryptic HDGFL2. A polyclonal goat antibody (gTEA1.2) against the C-terminal of the wild-type protein was used as detection antibody. This antibody was sulfo-tagged at a challenge ratio of 1:20 using the MSD GOLD SULFO-TAH NSH Ester Kit (Meso Scale Discovery LLC, Rockville, MD, USA), reaching a sulfo-tag label: protein conjugation ratio of 7.77:1. The antibody was diluted in MSD Diluent 100 at a concentration of 10µg/µl per well. Following incubation of the detection antibody, the plates were washed with PBS-T, and 150 µL of MSD GOLD Read Buffer A was added to each well. Plates were immediately measured on MESO QuickPlex SQ120 MM instrument. The limit of detection (LoD) for cryptic HDGFL2 was estimated by measuring 6Lwells/plate of diluent-only and calculating each signal as a ratio over the mean signal. The LoD was then calculated as the mean ratio plus two standard deviations. Following normalization of biofluid signals to the diluent-only signal, cryptic HDGFL2 peptide was considered ‘detectable’ only if sample signal ratios were higher than LoD. To compare measurements between plates, MSD signals were normalized to the diluent-only signal in their respective plates, with a ratio > 1 indicating an elevated cryptic HDGFL2 signal. These normalized values were then used to calculate cryptic HDGFL2 concentration based on the standard curve plate. The LoD for CSF:diluent-only and plasma:diluent-only were 1.05 and 1.10, respectively. Inter-assay CV was performed using 10 plasma samples from different patients. For these, two aliquots collected at the same time were assayed one month apart. MSD signal values were normalised as described before, and these ratios were used for inter-assay CV calculation.

### NfL quantification

Biofluid samples were assayed in duplicates and diluted 5-fold for CSF or 2-fold for plasma using Diluent 12 working solution (Meso Scale Discovery LLC, Rockville, MD, USA) to avoid hook effect and fit the assay signal into the linearity range. ELISA immunoassay was performed using the R-PLEX Human Neurofilament L Assays Kit (detection limit above 5.5 pg/ml) and MESO QuickPlex SQ120 MM instrument following the manufacturer’s instructions. Cut-off concentration was defined at 99% CI of the NfL levels in the control group.

### Statistical analysis

All the statistical analyses were performed using R Studio (version 4.3.3) and GraphPad Prism software (v9, Dotmatics, Boston, MA, USA). Since cryptic HDGFL2 signals are not normally distributed, analyses were performed using a non-parametric test. Group comparisons were conducted using the Kruskal-Wallis non-parametric test, with subsequent adjustment for multiple comparisons using the Benjamini-Hochberg false discovery rate (FDR) correction. Association between variables were tested using the Pearson correlation coefficient (r), which measures linear correlation in data sets. Receiver Operating Characteristic (ROC) analyses were performed using the “AUC” and “pROC” packages for R, to calculate sensitivity and specificity with relative 95% CI to evaluate the diagnostic accuracy of each biomarker in discriminating the main clinical groups. The data are presented as the meansL±Lstandard deviations (SD). Differences were considered statistically significant when *p-value*L<L0.05.

## RESULTS

### HDGFL2 cryptic peptide is increased in the CSF of individuals with TDP-43 proteinopathy

First, HDGFL2 cryptic peptide was measured to monitor TDP-43 loss of function in paired CSF and plasma samples from the DUH cohort (Figure 1A), using the MSD ELISA technology as previously described ^9^. Cryptic HDGFL2 signal was detected in CSF of ALS (37%), genetic FTD (29.4%) and bvFTD (31.25%) cases with suspected TDP-43 proteinopathy. Although it did not reach statistical significance in any of the groups, we observed a clear trend of increase in the ALS group compared with the healthy control group (*p*=0.092) (Figure 1B). The segregation of CSF samples based on sex showed a significant increase in HDGFL2 cryptic peptide signal in female ALS individuals compared to controls (p = 0.0191), but not in males (Figure 1C). The highest HDGFL2 signal values in the ALS female group (HDGFL2: diluent-only ratios of 2.78 and 1.93) correspond to carriers of ARPP21 missense variant c.1586C>T (Table S1). In contrast, cryptic HDGFL2 peptide signal can only be detected in the plasma of a few cases of ALS (10.34%), gFTD (17.65%) and bvFTD (12.5%) (Figure 1D). This measurement included two additional unpaired plasma samples in the ALS group (n=29) that were positive for the *ARPP21* missense variant c.1586C>T. However, in plasma, none of them were positive for cryptic HDFGL2. It should be noted that all positive plasma samples in the gFTD group were females (Figure 1E). Negative results in plasma may be due to the lack of correlation of cryptic HDGFL2 peptide signal between both biofluids (Figure S1A). Indeed, the cryptic concentration was 6-7-fold higher in CSF than in plasma (Figure S1B). Moreover, we also investigated the diagnostic and stratification accuracy of CSF HDGFL2 cryptic peptide to separate cognitively normal individuals from ALS and FTD patients. ROC analyses demonstrate a moderate accuracy in separating ALS and FTD (combination of bvFTD and genetic cases) from controls (AUC=0.68 and AUC=0.66, respectively) with higher specificity (80%) than sensitivity (58%) (Figure 1F).

**Figure 1.**
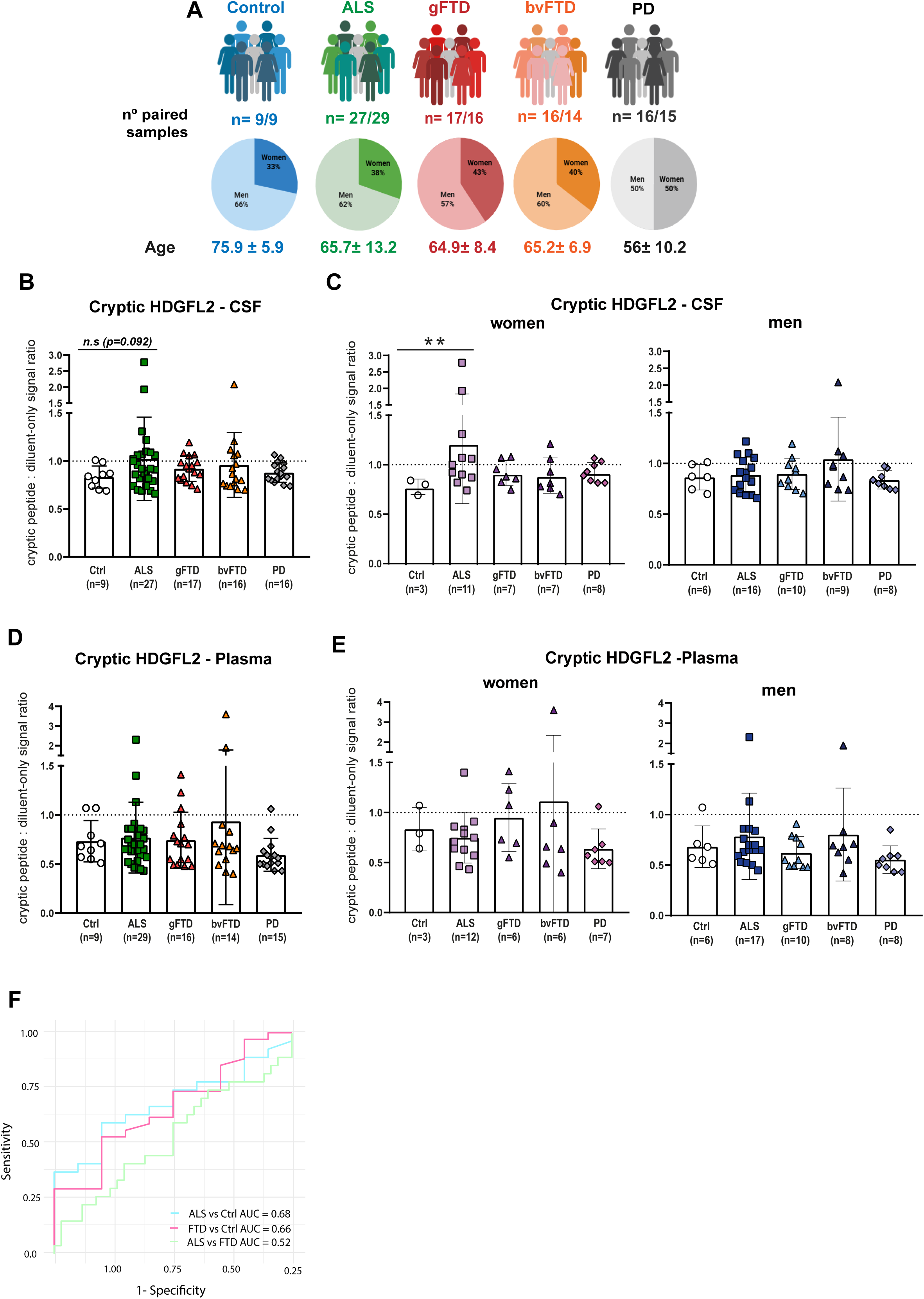
Cryptic HDGFL2 peptide detection in CSF and plasma samples from the DUH cohort. Graphical summary of the cases included from Donostia University Hospital (DUH), including controls (n=9), ALS (n=29), genetic FTD (gFTD, n=17), behavioral variant FTD (bvFTD, n=16) and LRRK2^+^ Parkinsońs disease (n=16) (A). Cryptic HDGFL2 peptide: diluent only signal ratios measured in CSF of all samples (B) and segregated in female and male (C) cases. Cryptic HDGFL2 peptide:diluent only signal ratios measured in plasma of all samples (D) and segregated in females and males (E) cases. ROC curve to assess the sensitivity and specificity of cryptic HDGFL2 as a diagnostic and prognostic biomarker in CSF samples from the DUH cohort (F). Data are expressed as mean ± SD. Statistical analysis was performed using the Kruskal-Wallis test, followed by Benjamini and Hochberg FDR correction for multiple comparisons; \*\**p*<0.01 vs control.

#### The level of NfL is increased in the CSF of individuals with TDP-43 proteinopathy

Levels of NfL in the DUH cohort were measured using MSD ELISA technology. We found an increase in NfL levels in CSF of ALS (81.48%), bvFTD (75%) and gFTD (64.7%) cases in comparison to the controls, whereas in plasma, only significant differences were found in ALS cases (*p*=0.0021) (Figure 2A and 2C). The segregation of the samples based on sex revealed an increase of NfL levels in CSF for both male and female ALS patients compared to healthy controls (*p*=0.0249 and *p*=0.0029), as well as for gFTD cases (*p*=0.0495) (Figure 2B). NfL levels in plasma were only significantly increased in male ALS cases compared to controls (*p*=0.0136), but not in females (Figure 2D). Levels of NfL did show a significant positive correlation (*p* < 0.001) between both biofluids (Figure S1C-S1D). As expected, NfL exhibited a high value for the discrimination of control *versus* ALS cases (AUC=0.89; 92% specificity; 91% sensitivity) in CSF, whereas its power is moderate for the discrimination of control *versus* FTD cases (AUC=0.67; 76% specificity; 59% sensitivity) and ALS *versus* FTD (AUC=0.77; 92% specificity; 58% sensitivity) (Figure 2E). Taken together, these results indicate that HDGFL2 cryptic peptide can be detected in the CSF of TDP-43 proteinopathy cases and is particularly high in female ALS patients, but with a modest discriminatory value. In contrast, NfL appears to be elevated in ALS, bvFTD and gFTD cases and has better predictive power than cryptic HDGFL2 peptide, but is not a biomarker specific for TDP-43 proteinopathies.

**Figure 2.**
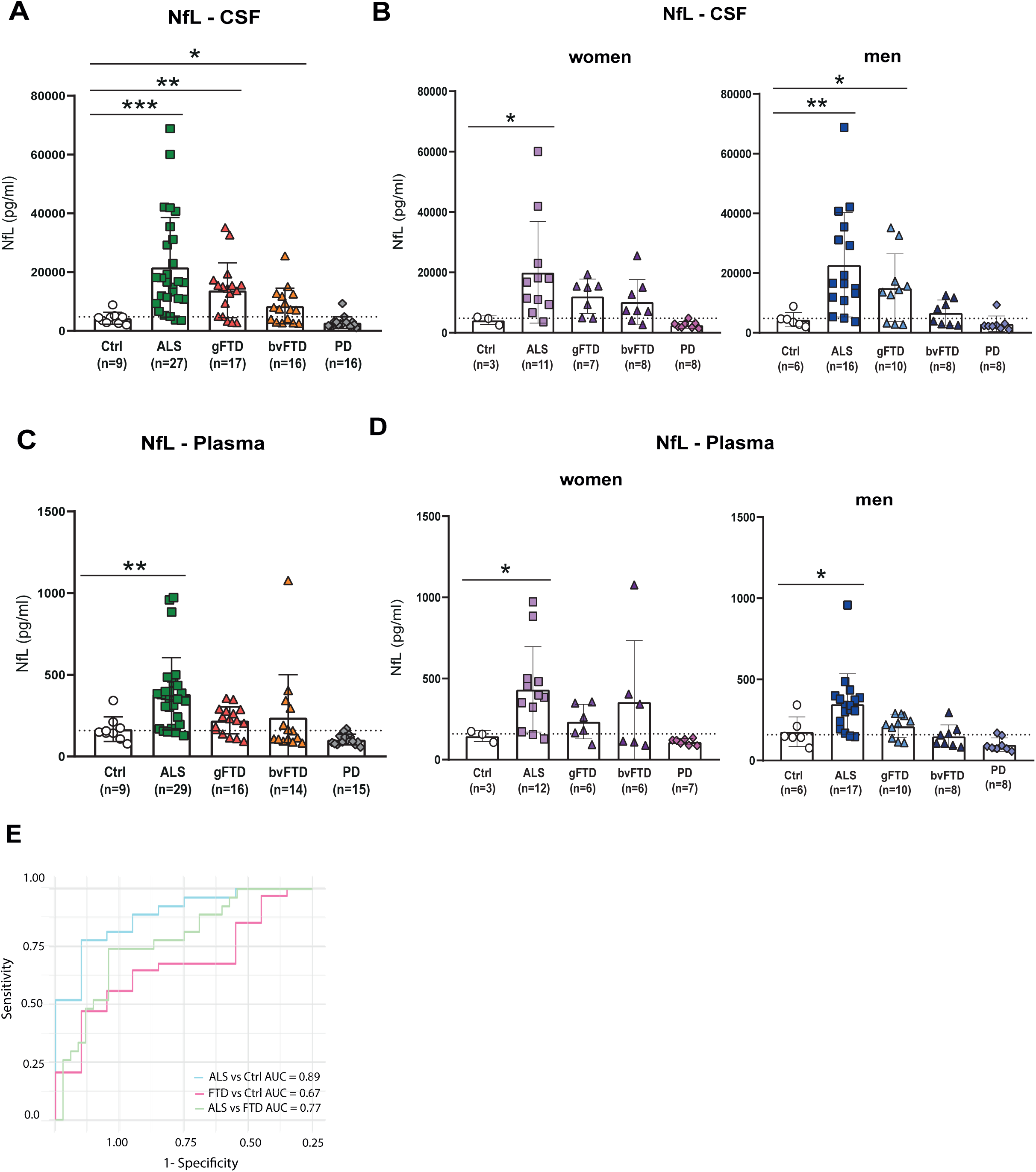
NfL levels are increased in CSF and plasma samples from the DUH cohort cases positive for TDP-43 proteinopathy. NfL concentration (pg/ml) measured in CSF of all samples (A) and segregated in female and male (B) cases. NfL concentration (pg/ml) measured in plasma of all samples (C) and segregated in females and males (D) cases. ROC curve to assess the sensitivity and specificity of cryptic HDGFL2 as a diagnostic and prognostic biomarker in CSF samples from the DUH cohort (E). Data are expressed as mean ± SD. Statistical analysis was performed using the Kruskal-Wallis test, followed by Benjamini and Hochberg FDR correction for multiple comparisons; \**p*<0.05; \*\**p*<0.01, \*\*\**p*<0.001 vs control.

#### NfL to HDGFL2 cryptic peptide ratio as a biomarker for TDP-43 proteinopathies

NfL is widely used in clinical practice as a neurodegenerative biomarker, but its broad profile limits its ability to exclude alternative diagnoses. The combination of NfL with HDGFL2 as a specific tracker of TDP-43 loss of function would provide a dual biomarker approach to improve diagnostic accuracy and prognostic capability for ALS and FTD patients. In this context, we calculated the ratio of NfL: HDGFL2 cryptic peptide levels. Interestingly, we found an increase of NfL: HDGFL2 ratio in both biofluids (CSF and plasma) of ALS patients (*p*=0.0002 and *p*=0.0019, respectively), as well as in CSF samples of gFTD cases compared to controls (*p*=0.0019), but not for bvFTD (which can present either TDP-43, Tau or even FUS pathology) (Figure 3A-D). These results highlight the value of combining HDGFL2 cryptic peptide and NfL as a dual biomarker strategy for use not only in patient diagnosis and follow-up but also in the stratification of FTLD-TDP from FTLD-Tau. Moreover, the combination of both biomarkers in CSF exhibited the highest value observed in this study for the discrimination of an ALS case *versus* a healthy control (AUC=0.91; 92% specificity; 80% sensitivity). NfL: HDGFL2 ratio also achieved higher values for the discrimination of FTD cases *versus* control (combination of bvFTD and gFTD cases) (AUC=0.76; 100% specificity; 59% sensitivity) than each of the biormarkers separately (Figure 3E). Finally, we also assessed the relationship between both biomarkers and the disease progression rate (DPR) in the ALS group. No significant correlation was found for HDGFL2 cryptic peptide (Figure 3F). Although the NfL concentration significantly increases with disease progression (*p*=0.0113) (Figure 3G), the best correlation in ALS cases is shown between the NfL: HDGFL2 cryptic peptide ratio and DPR in this group of patients (*p*=0.0026) (Figure 3H).

**Figure 3.**
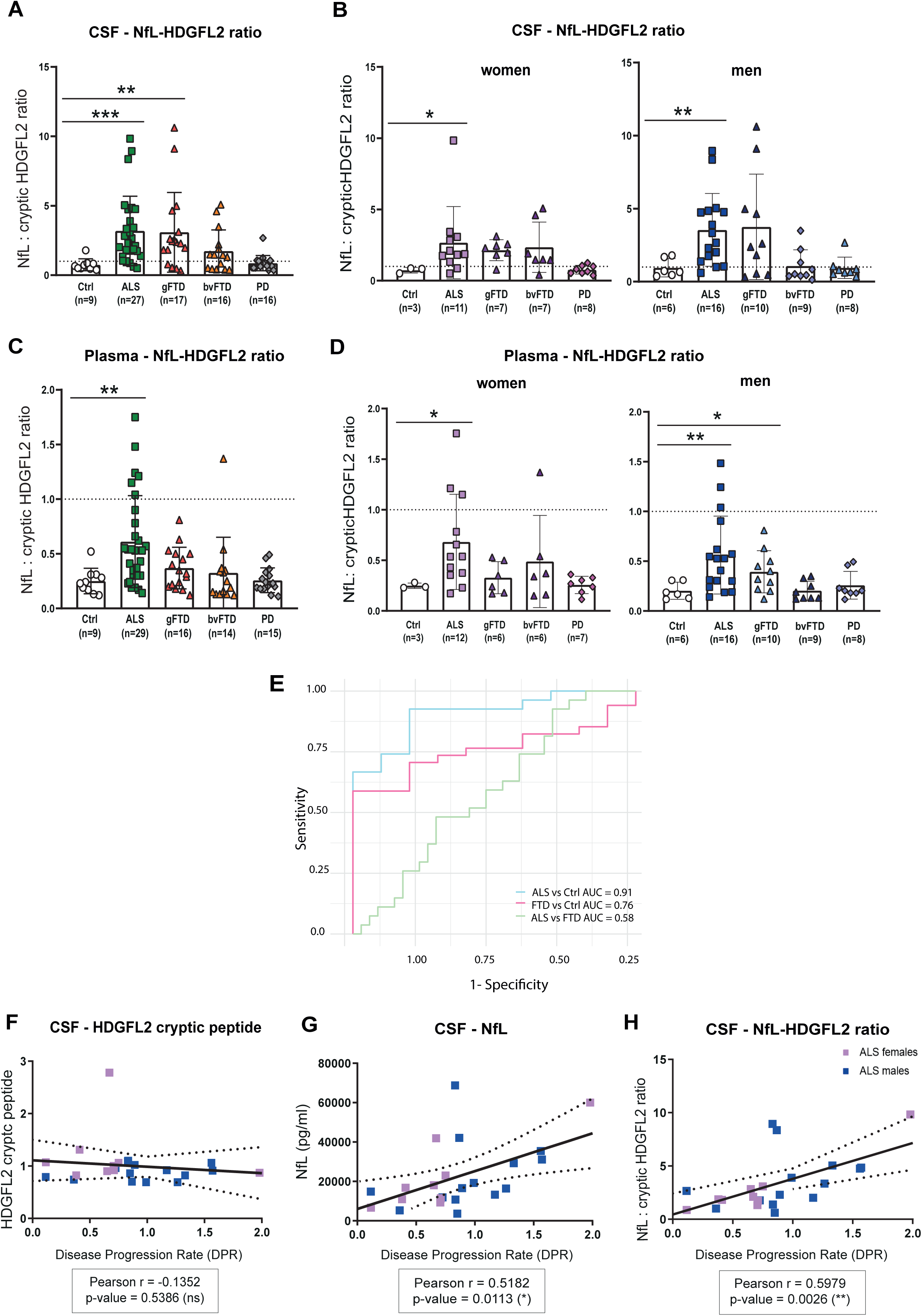
The ratio between NfL and cryptic HDGFL2 peptide showed a high diagnostic and prognostic power in ALS patients. NfL: HDGFL2 ratio measured in CSF of all samples (A) and segregated in female and male (B) cases. NfL: HDGFL2 ratio measured in plasma of all samples (C) and segregated in females and males (D) cases. ROC curve to assess the sensitivity and specificity of cryptic HDGFL2 as a diagnostic and prognostic biomarker in CSF samples from the DUH cohort (E). Pearson correlation two-tailed analysis to assess the linear relationship between levels of each biomarker in CSF - cryptic HDGFL2 (F), NfL (G) and NfL: HDGFL2 cryptic peptide ratio (H) - and disease progression rate (DPR) including only ALS cases (n=27). The correlation coefficient (r) indicates the strength and direction of the association. Data are expressed as mean ± SD. Statistical analysis was performed using the Kruskal-Wallis test, followed by Benjamini and Hochberg FDR correction for multiple comparisons; \**p*<0.05; \*\**p*<0.01, \*\*\**p*<0.001 vs control.

## DISCUSSION

TDP-43 nuclear depletion leads to the inclusion of CEs that result in the production of novel peptides ^5,8^. Recent studies have focused on a CE-derived peptide from HDGFL2 gene, which is detected in *post-mortem* tissue and correlates with phosphorylated TDP-43 levels ^10^. It has also been investigated as a sensitive biomarker for TDP-43 loss-of-function in the CSF of both symptomatic and presymptomatic individuals with ALS and FTD linked to *C9Orf72* ^9^. According to previous results, our study in an independent cohort detected cryptic HDGFL2 in CSF samples of ALS, bvFTD, and gFTD cases suspected of TDP-43 proteinopathy, showing high specificity but a moderate discriminatory power. CSF cryptic HDGFL2 levels were significantly high in the female ALS group, including two carriers of the ARPP21 missense variant c.1586C>T ^12^. This suggests that TDP-43 proteinopathy is not only restricted to *C9Orf72* as described by Irwin K.E and colleagues, but can also occur in other ALS-associated genes.

In plasma, cryptic HDGFL2 was only detected in a few cases of ALS and FTD, an expected result considering that cryptic HDGFL2 concentration was higher in the CSF. Cryptic HDGFL2 is a surrogate marker of TDP-43 nuclear depletion in neural cells. Its direct diffusion from the brain parenchyma to the CSF may explain its higher concentration in this biofluid. In contrast, biomarker release into the periphery may be slower and dependent on the mechanism and the molecular weight of the molecule^17^. It could also be facilitated by the disruption of the BBB^18,19^, described in ALS cases, although not consistently in all patients ^20,21^. These aspects may explain the lack of correlation of HDGFL2 cryptic peptide between the two biofluids evaluated in our study.

Consistent with prior research, NfL levels are elevated in both CSF and plasma of ALS and FTD patients, showing a strong correlation between both biofluids. Multicentric studies and our data confirm that increased NfL correlates with the age of onset, disease progression rate or survival in ALS and FTD, making it useful for monitoring disease ^22–24^. However, since NfL directly reflects axonal damage, its specificity in differentiating ALS or FTD between them or from other neurodegenerative conditions is limited ^1, 25^.

In contrast, HDGFL2 cryptic peptide results from TDP-43 loss-of-function-mediated aberrant splicing, providing a disease-specific marker for ALS-FTD spectrum. Previous studies have attempted to directly measure TDP-43 levels in CSF and plasma samples of ALS patients; however, results vary widely ^26^, prompting the combination of TDP-43 (or biomarkers related to its function) with other molecules to improve diagnostic performance. In this context, the ratio of HDGFL2 cryptic peptide and NfL offers a combined approach to improve the diagnosis and stratification of ALS and genetic cases of FTD with a clear TDP-43 proteinopathy. We observed that the NfL: HDGFL2 cryptic peptide ratio in CSF is significantly increased in ALS and gFTD, but not in bvFTD. These differences may reflect underlying variations in TDP-43 neuropathology between the two diseases. Cytosolic TDP-43 protein aggregates are more abundant in spinal motor neurons than in neurons located in the prefrontal and motor cortex^27,28^. Indeed, substantial heterogeneity in TDP-43 loss of function and delocalization appears to be more pronounced in ALS, corresponding to higher levels of HDGFL2 cryptic peptide. Moreover, the differences observed between genetic and behavioral FTD cases may be attributable to the fact that approximately 50% of patients with the behavioral clinical variant exhibit Tau pathology rather than TDP-43 ^29^. In contrast, most of our gFTD cases are carriers of the *PGRN* c.709-1G>A mutation, which is directly linked to TDP-43 proteinopathy.

The combination of both biomarkers showed the highest specificity observed in the DUH cohort. In plasma, this combination is also highly significant for ALS cases (not observed with HDGFL2 only) compared to controls, presenting a potential, easy-to-collect biomarker for the diagnosis and follow-up of this group of patients. The performance of NfL: cryptic HDGFL2 ratio in our study is better than the combination of TDP-43 and NfL in CSF in a previous study, which yielded an AUC = 0.843 for the discrimination of ALS from a healthy control ^30^. Other studies have evaluated the combination of three biomarkers to improve the diagnosis and stratification of neurodegenerative diseases ^31^. In our case, we achieved a discrimination power of 0.91 (92% specificity; 80% sensitivity) for ALS *versus* control, combining only two biomarkers. The NfL: HDGFL2 ratio also correlates with disease progression rate in the ALS group, where it performs better than each of the biomarkers separately, highlighting its value not only for diagnosis but also for the clinical follow-up of the patients, as has been observed in the DPR data.

In conclusion, our study confirms and extends previous evidence of the diagnostic value of the cryptic HDGFL2 biomarker in TDP-43 proteinopathies, highlighting how the integration of fluid biomarkers can improve diagnostic accuracy between ALS and FTD. Although our sample size is small, we were able to reproduce previous results in a completely independent and clinically characterized patient cohort. Future studies that combine novel surrogate markers of TDP-43 mis-splicing with more sensitive antibodies and molecular assays in multicentre and longitudinal cohorts will contribute to describing a combined biomarker strategy that could be implemented in the clinic.

## Data Availability

All data produced in the present work are contained in the manuscript

## Author contribution

Conceptualization: I.S.G, A.L.M, P.C.W, L.B; Methodology: K.E.I, J.P.L, P.C.W, I.S.G, P.G.A; Formal analysis: I.S.G, L.B; Resources: F.M.I, J.R.M, A.L.M, P.C.W, L.B; Data curation: I.S.G, P.G.A; Writing-original draft: I.S.G, L.B; Writing−review and editing: all authors; Supervision: P.C.W, L.B; Funding acquisition: L.B

## Consent for publication

All authors have read and agreed to the published version of the manuscript

## Ethics declaration

This study was approved by the Euskadi Medicine Research Ethics Committee (CEIm-E) PI+CES-BIOEF 2023-11. All participants gave informed consent for sample collection and participation in the study.

## Acknowledgements

We are grateful to patients and families for their collaboration. We also want to acknowledge Olatz Arnold and Jone López-Erauskin for their initial involvement in the development of this study and all members of the ‘NeuroRNA - Neurogenetics, RNA biology and therapies’ group for critical discussion of this work, as well as the Basque Biobank for providing CSF and plasma samples.

---

**Figure S1. Correlation of HDGFL2 cryptic peptide signal and NfL (pg/ml) in CSF and plasma samples**. Pearson correlation two-tailed analysis to assess the linear relationship between cryptic HDGFL2 (A) and NfL (C) in both biofluids, including ALS (n=27), genetic FTD (gFTD, n=17) 427 and behavioral variant FTD (bvFTD, n=16) cases. The correlation coefficient (r) indicates the strength and direction of association. Concentration of HDGFL2 cryptic peptide (ng/ml) (B) and NfL (pg/ml) (D) found in CSF and plasma samples. Statistical analysis was performed by Mann-Whitney U test for categorical variables; ***p<0.001 vs CSF concentration for each disease group.

