## Supplementary figures and images for "Light Chain Neurofilament to HDGFL2 cryptic peptide ratio as a fluid biomarker to monitor TDP-43 dysfunction in ALS and FTD"

### Supplementary Figure 1

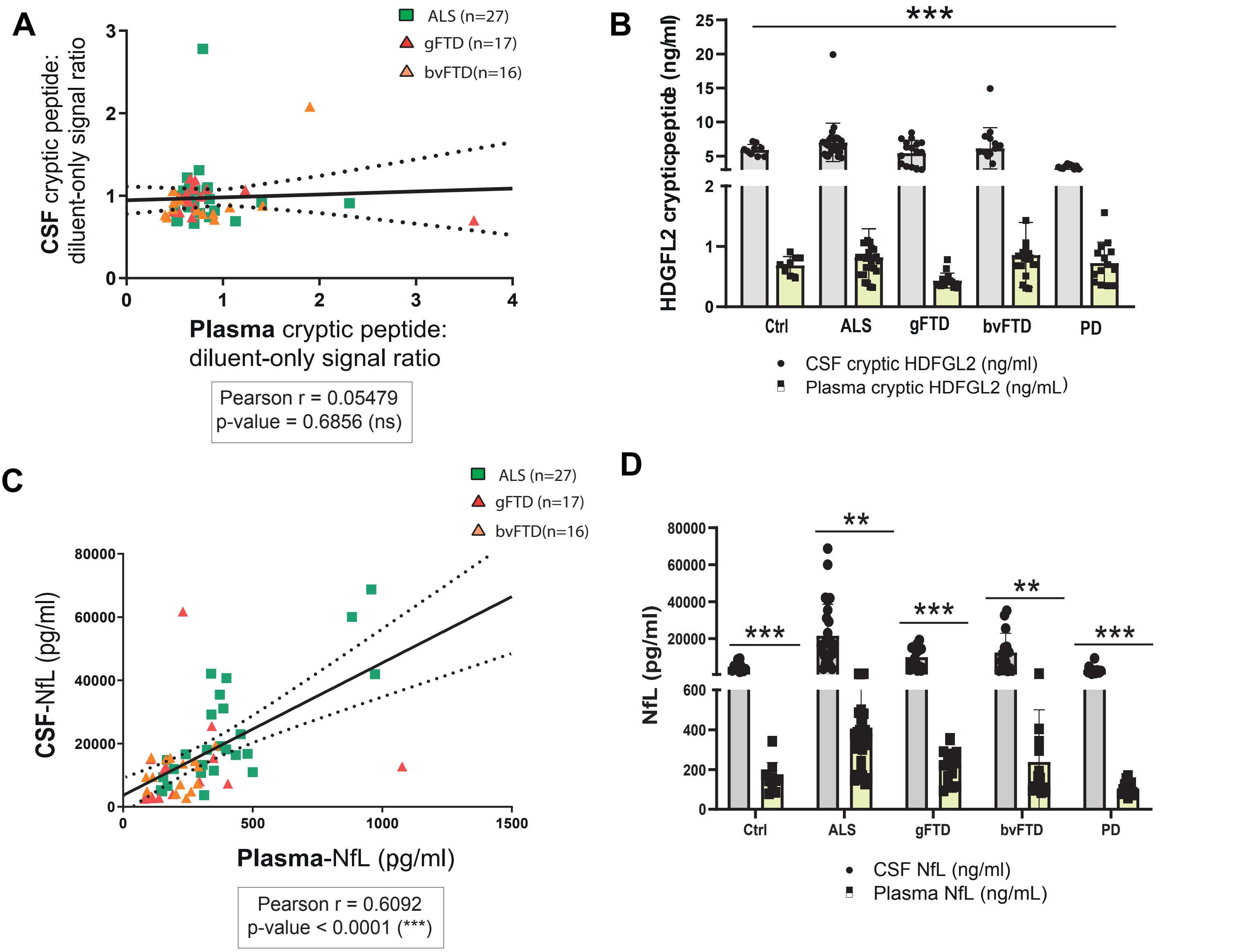
